# CD4+ and CD8+ T cell and antibody correlates of protection against Delta vaccine breakthrough infection: A nested case-control study within the PITCH study

**DOI:** 10.1101/2023.02.16.23285748

**Authors:** Isabel Neale, Mohammad Ali, Barbara Kronsteiner, Stephanie Longet, Priyanka Abraham, Alexandra S. Deeks, Anthony Brown, Shona C. Moore, Lizzie Stafford, Susan L. Dobson, Megan Plowright, Thomas A.H. Newman, Mary Y Wu, Crick COVID Immunity Pipeline, Edward J Carr, Rupert Beale, Ashley D Otter, Susan Hopkins, Victoria Hall, Adriana Tomic, Rebecca P. Payne, Eleanor Barnes, Alex Richter, Christopher J.A. Duncan, Lance Turtle, Thushan I. de Silva, Miles Carroll, Teresa Lambe, Paul Klenerman, Susanna Dunachie, the PITCH Consortium

**Author notes:** Corresponding authors: Susanna Dunachie, Peter Medawar Building for Pathogen Research, University of Oxford, OX1 3SY, UK., Paul Klenerman, Peter Medawar Building for Pathogen Research, University of Oxford, Oxford OX1 3SY, UK. Senior author (Teresa Lambe, Paul Klenerman, Susanna Dunachie). Secondary author list is included in the supplement.

## Abstract

T cell correlates of protection against SARS-CoV-2 infection after vaccination (‘vaccine breakthrough’) are incompletely defined, especially the specific contributions of CD4+ and CD8+ T cells. We studied 279 volunteers in the Protective Immunity from T Cells in Healthcare Workers (PITCH) UK study, including 32 cases (with SARS-CoV-2 positive testing after two vaccine doses during the Delta-dominant era) and 247 controls (no positive test nor anti-nucleocapsid seroconversion during this period). 28 days after second vaccination, before all breakthroughs occurred, cases had lower ancestral S- and RBD-specific immunoglobulin G titres and S1- and S2-specific T cell interferon gamma (IFNγ) responses compared with controls. In a subset of matched cases and controls, cases had lower CD4+ and CD8+ IFNγ and tumour necrosis factor responses to Delta S peptides with reduced CD8+ responses to Delta versus ancestral peptides compared with controls. Our findings support a protective role for T cells against Delta breakthrough infection.

## INTRODUCTION

Vaccination against severe acute respiratory syndrome coronavirus 2 (SARS-CoV-2) has been a crucial control strategy for the current COVID-19 pandemic. Although new SARS-CoV-2 variants continue to emerge and cause infection, vaccine protection against severe COVID-19 remains high for otherwise healthy individuals (Collie et al., 2022; Tartof et al., 2021). Nonetheless, infection or re-infection after a two-dose primary vaccination course, so-called ‘vaccine breakthrough’, occurred during the emergence and spread of variants of concern (VOC). As a result, booster (third) doses have been made available to all adults in many countries including the UK (UK Health Security Agency, 2022).

Understanding correlates of protection (CoP) against infection with SARS-CoV-2 could optimise delivery of current vaccines by informing boosting schedules and identifying individuals with suboptimal responses to vaccination. Identification of CoP could further facilitate licencing of modified or new vaccines based on immunogenicity and safety data rather than efficacy data (Feng et al., 2021), allowing new vaccines to more readily enter the marketplace.

Evidence supports a number of humoral correlates of protection against infection with SARS-CoV-2. Neutralising antibody (NAb) titres after vaccination correlate with vaccine efficacy and protection from infection in trials (Earle et al., 2021; Feng et al., 2021; Gilbert et al., 2022; Khoury et al., 2021) and NAbs from prior infection with ancestral variant are associated with protection against infection with ancestral (Addetia et al., 2020) and Alpha (Atti et al., 2022) variants. NAbs against SARS-CoV-2 are lower in the peri-infection period in those who experience Alpha vaccine breakthrough (Bergwerk et al., 2021), and are lower at diagnosis in those who experience Delta vaccine breakthrough (Chau et al., 2021) compared with uninfected controls. Anti-ancestral spike (S) and receptor binding domain (RBD) immunoglobulin G (IgG) titres also correlate with protection from infection with the Alpha variant (Feng et al., 2021) and Delta variant (Aldridge et al., 2022; Smoot et al., 2022; Wei et al., 2022) after vaccination. Ancestral variant RBD-specific memory B cell responses were lower in Delta breakthrough cases at the point of diagnosis compared to uninfected close contacts (Tay et al., 2022). Levels of serum anti-Spike and anti-RBD IgA 2-4 weeks after second dose of BNT162b2 vaccine were also lower in those who experienced subsequent Alpha or Gamma vaccine breakthrough compared to those who remained uninfected (Sheikh-Mohamed et al., 2022).

T cells play an important role in control of SARS-CoV-2 infection and in reducing disease severity upon infection (Kedzierska and Thomas, 2022; Moss, 2022). In primary infection, strong SARS-CoV-2 specific CD4+ and CD8+ T cell responses are associated with reduced disease severity (Rydyznski Moderbacher et al., 2020). Several studies have demonstrated that spike-specific T cell responses are largely maintained against VOC (Gao et al., 2022; Moore et al., 2022; Payne et al., 2021; Skelly et al., 2021; Tarke et al., 2022), despite evasion of antibody responses (Cele et al., 2022; Planas et al., 2021; Zhou et al., 2021). This supports T cells contributing to the continued effectiveness of vaccines against severe disease.

However, identifying a clear role for T cells in preventing infection is difficult due to the lack of sensitive, scalable T cell assays, and potential confounding effects of antibody responses (Kedzierska and Thomas, 2022; Kent et al., 2022). Nonetheless, an increasing number of reports indicate that T cells are likely to be important in protection against SARS-CoV-2 infection. Unvaccinated individuals who are exposed to SARS-CoV-2 but remain antibody and PCR-negative show T cell responses (Ogbe et al., 2021), and further studies demonstrate that pre-existing, cross-reactive T cell responses are associated with protection from overt infection (Kundu et al., 2022; Swadling et al., 2022). In an unvaccinated cohort study, seronegative individuals with detectable SARS-CoV-2-specific T cell responses had a lower risk of subsequent infection than seronegative individuals without positive T cell responses (Molodtsov et al., 2022). More recently, higher whole-blood interferon gamma (IFNγ) responses to SARS-CoV-2 peptides among individuals who had received at least one vaccine dose were associated with a lower risk of breakthrough infection over the subsequent 6-month period (Scurr et al., 2022). There are also indications that the combination of cellular and humoral responses may be important in protecting against Delta and Omicron breakthrough infection. The combination of high NAb titre and high S1-specific IFNγ responses is associated with protection against breakthrough (Almendro-Vázquez et al., 2022). Also, ‘high responders’ to vaccination, characterised by high antibody, enriched CD4+ and CD8+ central memory 1, and durable CD8+ T cell responses, are more protected against symptomatic breakthrough compared to ‘low responders’, characterised by low antibody responses and more prevalent T effector memory 2 responses (Brasu et al., 2022). Although these data indicate an important role for T cells, it is still unclear to what extent T cell protection is mediated by CD4+ versus CD8+ T cells, as in many studies only total T cell responses are investigated. However, depletion of vaccine-induced CD8+ T cells in rhesus macaques and subsequent challenge demonstrated that CD8+ T cells contribute to vaccine protection against SARS-CoV-2 replication in this model (Liu et al., 2022). Therefore, further research is needed to define possible T cell CoP against SARS-CoV-2 infection, in particular the contributions of CD4+ and CD8+ T cells to protection in humans.

Large-scale prospective studies with recruitment and sampling of individuals prior to breakthrough give opportunity to better understand possible T cell CoP against SARS-CoV-2 (Kent et al., 2022). The Protective Immunity from T Cells in Healthcare workers (PITCH) consortium is a prospective longitudinal cohort study of cellular immune responses to vaccination and infection in healthcare workers (HCWs) in the UK (http://www.pitch-study.org/), aligned with the wider SARS-CoV-2 immunity & reinfection evaluation (SIREN) study (Hall et al., 2022; Hall et al., 2021a; Hall et al., 2021b). The PITCH cohort demonstrated that an extended interval between first and second dose of the BNT162b2 vaccine was associated with higher NAb and anti-S IgG binding titre and lower S-specific T cell response magnitude but higher interleukin-2 (IL-2) production compared to a shorter interval (Payne et al., 2021), findings which were also demonstrated in a separate study of older adults (Parry et al., 2022). The PITCH cohort includes individuals studied from April 2020 onwards, with data from pre-vaccination through to after the fourth vaccine dose, allowing sampling of well-characterised participants who subsequently experienced infection. The first vaccine breakthrough case in a PITCH participant was reported on the 10^th^ June 2021, at which time, Delta was the predominant variant circulating in the UK (UK Health Security Agency, 2021). A further 35 reports of vaccine breakthrough occurred in the cohort before the first Omicron cases were reported in the UK on 27^th^ November 2021 (Department of Health and Social Care, 2021). This offers an opportunity to use samples stored from before breakthrough infection to investigate CoP against infection after vaccination.

In the current study, we conducted a case-control study within the PITCH framework, using cases who reported infection during the Delta-dominant era in the UK, and controls who did not experience vaccine breakthrough during this time. Cellular and humoral immunity at the timepoint 28 days after the second vaccine dose (V2+28) was studied using an unmatched overall design in order to investigate whether peak post-vaccination responses are associated with subsequent breakthrough infection, irrespective of factors which may influence the level of response. A subset of cases and matched controls were selected for more detailed analysis of cellular immunity. We observed lower S- and RBD-specific IgG binding titre and lower S1- and S2-specific T cell responses by IFNγ ELISpot assay in cases compared with controls at V2+28. Using intracellular cytokine staining (ICS), we observed lower CD4+ and CD8+ IFNγ and tumour necrosis factor (TNF) responses to Delta peptides among cases compared with matched controls.

## RESULTS

### Breakthrough infections during the Delta predominant era in the UK

Between 10^th^ June and 27^th^ November 2021, there were 36 self-reports of vaccine breakthrough (positive polymerase chain reaction (PCR) or lateral flow device (LFD) test for SARS-CoV-2 >14 days after second vaccine dose and <7 days after third vaccine dose) in the PITCH cohort. These participants had been vaccinated with two doses of SARS-CoV-2 vaccine between 6^th^ January and 17^th^ July 2021. 26 (72.2%) participants received BNT162b2 (Pfizer/BioNTech) and 10 (27.8%) received AZD1222 (Oxford/AstraZeneca) (Table 1). The median age of vaccine breakthrough cases was 43 (interquartile range [IQR] 31.5-49; range 22-72), 30 (83.3%) cases were female, and the majority (93.3%) of cases were of white ethnicity, reflecting the demographics of the underlying SIREN and PITCH study cohorts (Hall et al., 2021b; Payne et al., 2021). The median time between second vaccine dose and positive PCR or LFD test was 156 days (IQR 123.8-180.5; range 48-253).

**Table 1.** Demographic characteristics of vaccine breakthrough cases compared to controls and factors associated with vaccine breakthrough in multivariable logistic regression analysis.

|  | All<br>(n=414) | Controls<br>(n=378) | Cases<br>(n=36) | OR<sup>1</sup> (95% CI) | p value | Adjusted OR<sup>2</sup><br>(95% CI) | p value |
| --- | --- | --- | --- | --- | --- | --- | --- |
| Age |  |  |  |  |  |  |  |
| 20-29 | 78 (18.8%) | 70 (18.5%) | 8 (22.2%) | 1 (ref) | 0.396 | 1 (ref) | 0.456 |
| 30-39 | 86 (20.8%) | 78 (20.6%) | 8 (22.2%) | 0.90 (0.31-2.56) |  | 0.99 (0.34-2.86) |  |
| 40-49 | 109 (26.3%) | 97 (25.7%) | 12 (33.3%) | 1.08 (0.43-2.89) |  | 1.26 (0.49-3.44) |  |
| 50-59 | 97 (23.4%) | 93 (24.6%) | 4 (11.1%) | 0.38 (0.10-1.24) |  | 0.45 (0.11-1.51) |  |
| 60+ | 44 (10.6%) | 40 (10.6%) | 4 (11.1%) | 0.88 (0.22-2.96) |  | 1.19 (0.30-4.20) |  |
| Sex |  |  |  |  |  |  |  |
| Female | 310 (74.9%) | 280 (74.1%) | 30 (83.3%) | 1 (ref) | 0.203 | 1 (ref) | 0.078 |
| Male | 104 (25.1%) | 98 (25.9%) | 6 (16.7%) | 0.57 (0.21-1.32) |  | 0.46 (0.17-1.08) |  |
| Ethnicity (self-reported) |  |  |  |  |  |  |  |
| White | 298 (84.4%) | 270 (83.6%) | 28 (93.3%) | 1 (ref) | 0.265 | - | - |
| Asian | 37 (10.5%) | 36 (11.1%) | 1 (3.3%) | 0.27 (0.02-1.32) |  | - |  |
| Other | 18 (5.1%) | 17 (5.3%) | 1 (3.3%) | 0.57 (0.03-2.93) |  | - |  |
| Unreported | 61 | 55 | 6 | - |  | - |  |
| BMI (kg/m<sup>2</sup>) |  |  |  |  |  |  |  |
| Not obese (<30) | 162 (89.5%) | 150 (89.8%) | 12 (85.7%) | 1 (ref) | 0.645 | - | - |
| Obese (≥30.0) | 19 (10.5%) | 17 (10.2%) | 2 (14.3%) | 1.47 (0.22-6.01) |  | - |  |
| Unreported | 233 | 211 | 22 | - |  | - |  |
| Vaccine type |  |  |  |  |  |  |  |
| BNT162b2 | 336 (81.2%) | 310 (82.0%) | 26 (72.2%) | 1 (ref) | 0.171 | - | - |
| AZD1222 | 78 (18.8%) | 68 (18.0%) | 10 (27.8%) | 1.75 (0.77-3.71) |  | - |  |
| Infection history |  |  |  |  |  |  |  |
| Naïve | 244 (58.9%) | 214 (56.6%) | 30 (83.3%) | 1 (ref) | 0.001 | 1 (ref) | 0.0009 |
| Previous SARS-CoV-2 | 170 (41.1%) | 164 (43.4%) | 6 (16.7%) | 0.26 (0.10-0.60) |  | 0.25 (0.09-0.58) |  |

<sup>1</sup>Crude odds ratio derived from single logistic regression.
<sup>2</sup>Adjusted odds ratio derived from multiple logistic regression. The final model included and adjusted for age, sex, and infection history.

### Vaccine breakthrough is more common in individuals without history of previous infection

As expected, previous SARS-CoV-2 infection contributed to protection against further infection. Comparison of demographic characteristics of all breakthrough infection cases (n=36) with controls, defined as eligible individuals in PITCH who did not report a positive PCR or LFD by, and were still enrolled at, the 27^th^ November 2021 or their third vaccine dose (n=378), showed that the odds of vaccine breakthrough was higher among those previously infection-naïve compared to those previously infected with SARS-CoV-2 before vaccination (p=0.0009, Table 1, Supplementary Figure S1). No significant association was found between odds of vaccine breakthrough infection and age, sex, body mass index (BMI), ethnicity or vaccine type. All cases sampled at V2+28 (n=32) were then selected for a case-control study with controls (n=247) being those who had been sampled at the V2+28 timepoint and who did not report vaccine breakthrough or show evidence of anti-nucleocapsid (N) IgG seroconversion during the period of interest (Figure 1A).

**Figure 1.**
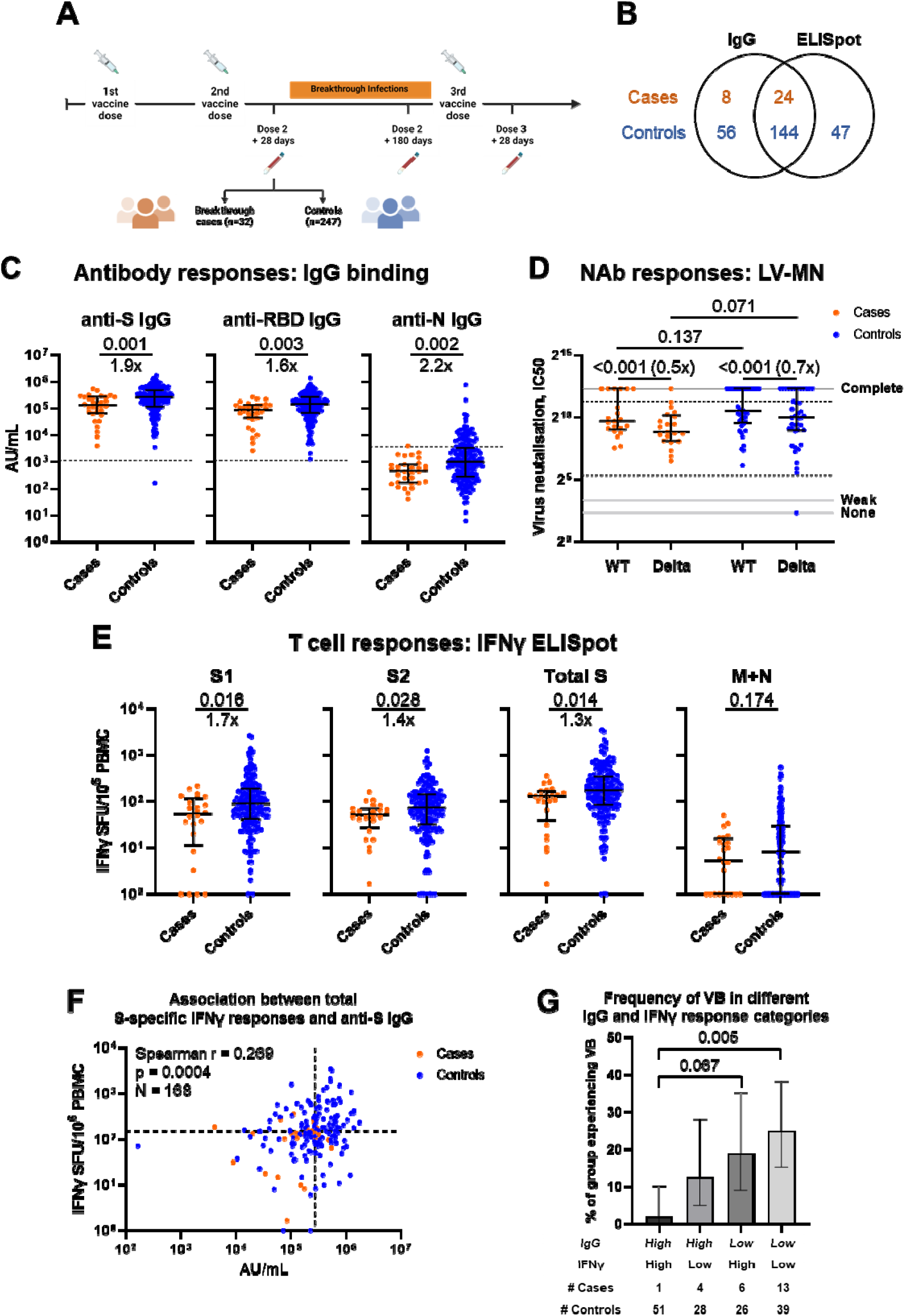
Study design and comparison of antibody and T cell responses between cases and controls at 28 days after second vaccine dose. **(A)** Schematic representation of study design including vaccination and breakthrough infection time points. Created using Biorender.com. **(B)** Venn diagram illustrating number of subjects with IgG and/or IFNγ ELISpot measurements, by case/control status. **(C)** Ancestral SARS-CoV-2 spike- (S), receptor binding domain- (RBD) and nucleoprotein- (N) specific IgG binding titre in cases (n=32) and controls (n=200), as measured by MSD. Dashed lines represent threshold for positive response (SARS-CoV-2 S IgG 1160 AU/mL, RDB IgG 1169 AU/mL, N IgG 3874 AU/mL). **(D)** Neutralising antibody (NAb) titres against ancestral (WT) and Delta (B1.617.2) isolates in a subset of cases (n=21) and controls (n=41) measured by live virus microneutralisation (LV-MN) assay. Area between IC50=40 and IC50=2560 (dashed black lines) corresponds to the quantitative range of the assay. IC50=5, IC50=10 and IC50=5120 (solid grey lines) represent no, weak and complete inhibition respectively. **(E)** T cell responses to peptide pools representing ancestral SARS-CoV-2 S1, S2, total S (summation of S1 and S2 responses) and membrane (M) and nucleoprotein (N) responses in cases (n=24) and controls (n=191), as measured by IFNγ ELISpot assay. **(F)** Correlation between anti-S IgG titre, as measured by MSD, and total S T cell responses, as measured by IFNγ ELISpot, for all cases (n=24) and controls (n=144) where both parameters were measured. Dashed lines represent median value for each parameter. **(G)** Frequency of individuals with vaccine breakthrough (VB) in each quadrant of the graph in (F). Individuals are classified as having ‘High’ or ‘Low’ IgG and IFNγ responses on the basis of whether their response is above or below median for each variable. Number of cases and controls per group are shown below the graph. Values of 0 are replaced with 1 for representation on the logarithmic scale. Orange circles represent cases, blue circles represent controls. Bars represent median of each group (C-E). Error bars represent interquartile range (C-E) or 95% confidence interval (G). Two-tailed p-values derived from Mann-Whitney U tests (comparing cases to controls, (C-E)), Wilcoxon matched pairs signed rank tests (comparing WT and Delta responses, (D)) or pairwise Fisher’s exact tests (G) shown above linking lines. For clarity, only adjusted p values <0.1 are shown in (G). Fold change in median response shown below linking lines or in brackets, calculated as median response for controls divided by median response for cases, or median response for Delta divided by median response for WT.

### Cases have lower humoral and cellular responses than controls at V2+28

Comparing all cases with a V2+28 sample to all available controls, cases had lower anti-S (p=0.001), -RBD (p=0.003) and -N (p=0.002) IgG binding titre compared with controls (Figure 1B, C). In a subset of cases (n=21) and controls (n=41), NAb titre against ancestral and Delta variants was measured (Figure 1D). No difference in median NAb titre against ancestral variant was detected between cases and controls, although the sample size was small. There was a trend for lower median NAb titre against the Delta variant among cases compared to controls (p=0.071). Both cases and controls displayed a reduction in NAb titre to Delta compared to ancestral virus (both p<0.001). Cases also demonstrated lower ancestral S1- (p=0.016), S2- (p=0.028) and total S- (p=0.014) specific IFNγ ELISpot responses compared to controls at the V2+28 timepoint (Figure 1E). Previous infection was a strong determinant of both IgG and IFNγ responses (Supplementary Figure S2 A-G). In a subset of individuals where cell availability permitted, IFNγ responses to peptide pools representing Delta S1 and S2 were assessed (Supplementary Figure S2 H-J). However, no difference was detected in responses between cases and controls, possibly due to the small sample size in this analysis.

There was weak correlation between individual anti-S IgG titre and total S IFNγ responses (Figure 1F). To investigate the combined effect of antibody and cellular responses on the risk of vaccine breakthrough, individuals were categorised as having ‘High’ or ‘Low’ anti-S IgG and S-specific IFNγ responses on the basis of whether the response was above or below the median, such that individuals were divided into four categories (Figure 1G). The proportion of individuals experiencing vaccine breakthrough was significantly different between categories (Fisher’s exact test, p=0.003). The proportion of those experiencing vaccine breakthrough was lowest among those with High IgG and High IFNγ responses, significantly lower than those with Low IgG and Low IFNγ responses (pairwise Fisher’s exact test, adjusted p=0.005). Those with either High IgG and Low IFNγ responses or Low IgG and High IFNγ responses had intermediate risk of breakthrough, although there was a trend for a difference between those with Low IgG and High IFNγ responses and those with High IgG and High IFNγ responses (adjusted p=0.067), highlighting the importance of the IgG response.

### Cases have lower S-specific T cell responses to Delta peptides compared to controls at V2+28

In a subset of cases and matched controls, all of whom were SARS-CoV-2 infection-naïve at vaccination, further analysis of cellular CoP was undertaken. Ancestral and Delta S-specific memory B cell frequency was compared in a subset of cases (n=10) and matched controls (n=11) where peripheral blood mononuclear cell (PBMC) samples were available (Figure 2A, B). There was a trend for lower Delta S-specific memory B cell frequency among cases compared to controls (p=0.088). However, there was high variation among controls and an outlier among cases, meaning that these data do not allow a conclusion to be drawn.

**Figure 2.**
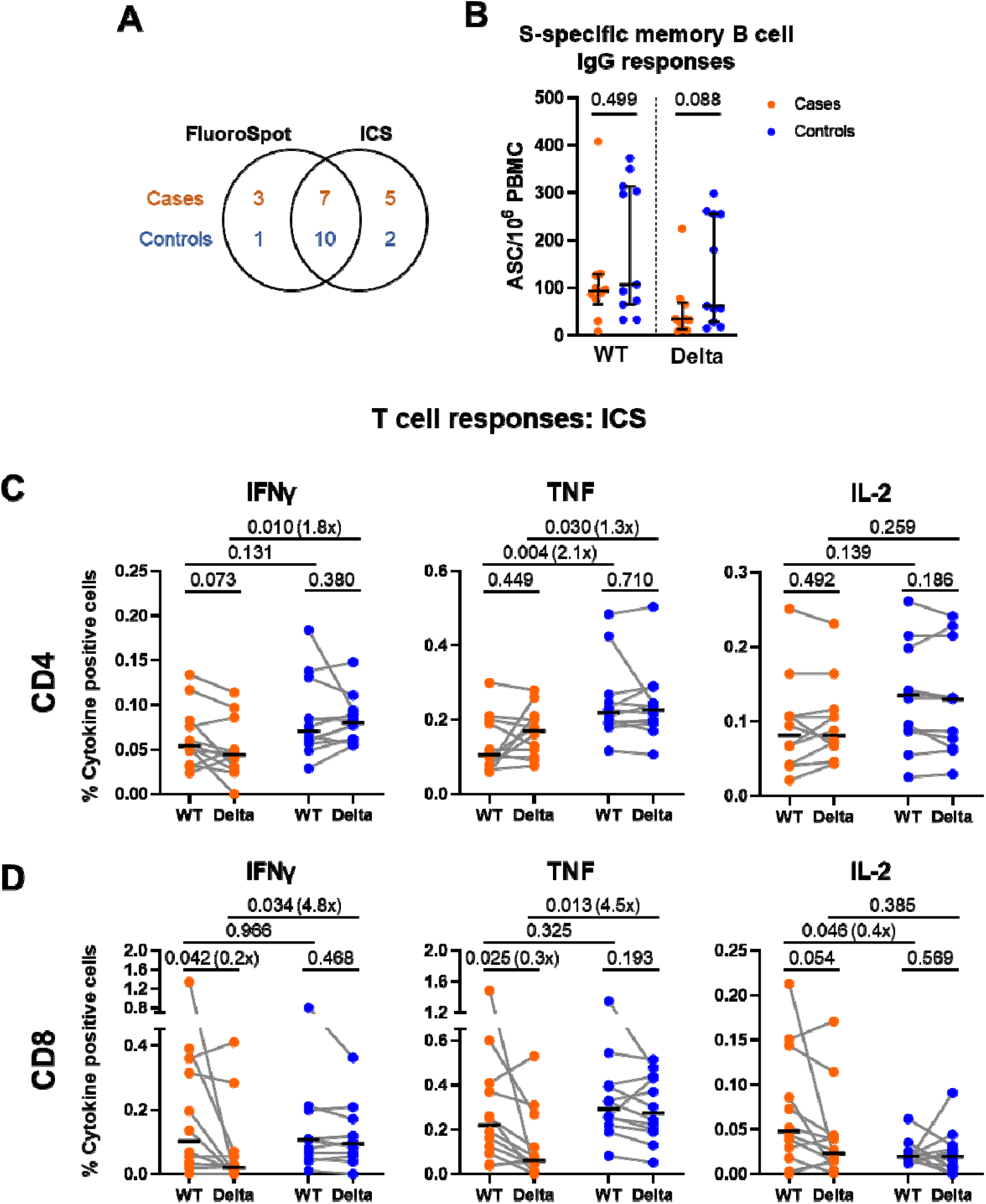
Comparison of memory B cell and T cell responses in a subset of cases and matched controls at 28 days after second vaccine dose. **(A)** Venn diagram illustrating the overlapping subsets of cases and controls analysed in (B-D). **(B)** Memory B cell responses against ancestral (WT) and Delta (B.1.617.2; AY.1, AY.2, AY.3) SARS-CoV-2 spike (S) in a subset of cases (n=10) and matched controls (n=11) measured by memory B cell IgG FluoroSpot. **(C)** Proportion of cells expressing IFNγ, TNF or IL-2 in response to peptide pools representing ancestral (WT) or Delta (B.1.617.2) spike in the CD4+ T cell population and **(D)** CD8+ T cell population, in a subset of cases and matched controls (n=12 per group), as measured by intracellular cytokine staining (ICS) and flow cytometry. Populations were analysed by gating on single, live, CD3+ cells. Orange circles represent cases, blue circles represent controls. Bars represent median of each group. Error bars represent interquartile range. Paired data from the same individual are linked by grey lines. Two-tailed p-values derived from Mann-Whitney U tests (comparing cases to controls) or Wilcoxon matched pairs signed rank tests (comparing WT and Delta responses) shown above linking lines. Fold change in median response shown in brackets for comparisons where p<0.05, calculated as median response for controls divided by median response for cases, or median response for Delta divided by median response for WT.

CD4+ and CD8+ T cell responses were further investigated by intracellular cytokine staining (ICS) in a subset of cases and matched controls where PBMC were available (n=12 per group). For CD4+ responses, there was a trend for lower frequency of IFNγ and TNF expressing cells in cases compared with controls, for both ancestral and Delta S peptide pools (Figure 2C). This difference was significant for TNF in response to ancestral S (p=0.004), and IFNγ (p=0.010) and TNF (p=0.030) in response to Delta S peptides. There was high variability in frequency of IL-2 expressing CD4+ T cells among cases and controls, with no difference detected in median frequency between groups. Across CD8+ T cell responses, there was a similar pattern, with a difference detected in frequency of IFNγ (p=0.034) and TNF (p=0.013) expressing cells in response to Delta S peptides (Figure 2D). This difference appeared to be driven by a reduced response among cases to Delta peptides compared to ancestral peptides, which was not present in controls. CD8+ IL-2 responses appeared to be slightly higher in cases compared to controls in response to ancestral S (Figure 2D, p=0.046). However, overall CD8+ IL-2 responses were very low and this trend appeared to be driven by one outlier with very high CD8+ responses across all cytokines.

Across IFNγ and TNF responses, on average over half of the response was contributed by CD4+ T cells, however there was substantial variation between individuals (Supplementary Figure S3A, B). A higher proportion of the TNF response to Delta peptide pools was contributed by CD4+ T cells among cases compared to controls (p=0.028). CD4+ T cells contributed almost all of the IL-2 response to ancestral and Delta S peptide pools across the majority of cases and controls (Supplementary Figure S3C). Polyfunctionality analysis indicated that cases had a lower frequency of TNF/IFNγ+ double positive CD4+ and CD8+ T cells in response to Delta peptides compared with controls (Supplementary Figure S3E, G). There was also a trend for lower frequency of TNF+ single positive CD4+ and CD8+ T cells among cases compared to controls (Supplementary Figure S3 D-G).

## DISCUSSION

This study examines potential CoP against SARS-CoV-2 breakthrough infection after two vaccine doses during the Delta-predominant era in the UK. Breakthrough infection was less likely in those with previous SARS-CoV-2 infection compared with those previously infection-naïve, consistent with other reports (Dhumal et al., 2022; Torres et al., 2022a). Cases showed lower anti-S and -RBD IgG binding titre, as well as lower S1- and S2-specific IFNγ T cell responses by ELISpot assay, compared with controls. In further analysis by ICS, lower CD4+ and CD8+ IFNγ and TNF responses to Delta S peptide pools were found among cases compared to controls. Our detailed look at spike-specific T cell function with sensitive assays provides further evidence of T cell CoP to add to established serological CoP from large studies of humoral function.

Our finding that anti-S and -RBD IgG binding titres differ between those who go on to experience breakthrough and those who do not experience overt infection is consistent with a number of other reports which previously demonstrated an association between these parameters and risk of breakthrough infection (Aldridge et al., 2022; Feng et al., 2021; Wei et al., 2022). The SIREN study is also well placed to look at serological CoP against reinfection in a large cohort of HCWs which includes several cases and controls from this study. SIREN recently reported that anti-S antibody levels and NAb titres correlated with protection against reinfection during the Alpha wave, prior to vaccination (Atti et al., 2022).

In this study, vaccine breakthrough was associated with lower IFNγ T cell responses by ELISpot, and lower CD4+ and CD8+ IFNγ and TNF responses to Delta peptides in ICS, even using a modest sample size. There are increasing reports indicating an important role for T cell CoP alongside serological CoP (Almendro-Vázquez et al., 2022; Molodtsov et al., 2022; Scurr et al., 2022). Our findings using the IFNγ ELISpot assay help to validate these reports. Our more detailed findings using ICS support a role for both CD4+ and CD8+ T cells in protecting against susceptibility to overt infection with the Delta variant. The apparent drop in CD8+ IFNγ and TNF responses to Delta peptides compared with ancestral peptides among cases was particularly striking and suggests a role for variant-specific CD8+ T cells in protection against infection with the Delta variant. Previous studies in a rhesus macaque model support a role for CD8+ T cell responses in protection against re-challenge in the context of waning antibody levels (McMahan et al., 2021), and a direct role in vaccine-induced protection against SARS-CoV-2 replication (Liu et al., 2022). In addition, infection-naïve vaccinees with high antibody responses have enriched CD4+ and CD8+ central memory 1 cell responses and durable CD8+ T cell responses to ancestral and VOC S peptides compared to those with low antibody responses, and are protected against symptomatic breakthrough infection with Delta or Omicron (Brasu et al., 2022). Virus-specific CD8+ T cells are associated with protection against infection with other viral pathogens. For example, in the context of influenza A virus (IAV) epidemics, each fold increase in pre-existing H3N2- or seasonal H1N1-specific CD8+ T cell response is associated with reduced odds of infection with H3N2 or pandemic H1N1 IAV respectively (Tsang et al., 2022). IFNγ+IL2-CD8+ T cells specific for conserved IAV core protein epitopes are also associated with protection against symptomatic disease caused by pandemic H1N1 IAV (Sridhar et al., 2013). Several human studies investigating immune responses around the time of SARS-CoV-2 breakthrough infection have not identified differences in T cell responses between cases and controls, but have identified differences in terms of NAb titre (Park et al., 2022; Rovida et al., 2021) and memory B cell responses (Tay et al., 2022). The difference in results between these studies and our study may be due to different timing of sampling and/or SARS-CoV-2 variant.

Our study focused on the Delta wave of infections in the UK, during a time at which the study participants had received two vaccine doses. However, the Delta variant has been largely displaced by Omicron in the UK and globally, and in the UK a third SARS-CoV-2 vaccine dose has been recommended to all adults (UK Health Security Agency, 2022) with further doses for those at increased risk and HCWs. The generalisability of the findings of this study to Omicron breakthroughs occurring after two or three vaccine doses needs further evaluation. Studies of CoP against Omicron have reported mixed results. A study investigating breakthrough after two vaccine doses found that there was no difference in anti-RBD titre between cases who subsequently experienced Omicron breakthrough and controls (Smoot et al., 2022). Other studies did not identify differences in anti-S IgG titre (Stærke et al., 2022), anti-RBD IgG titre, Omicron NAb titre or T cell IFNγ responses (Torres et al., 2022b) after a third vaccine dose between those who subsequently experienced Omicron breakthrough and those who did not. However, others have reported that low anti-S Ab titre and weak neutralisation post-third dose is associated with higher risk of breakthrough (Mohlendick et al., 2022). In the peri-infection period, low pre-infection anti-S IgG titre is associated with breakthrough (Nunes et al., 2022); however, it has also been shown that Omicron breakthrough can occur in those with high NAb titres (Roeder et al., 2022).

## Limitations

Important limitations are as follows. (i) The sample size for cases is relatively small, despite follow up of a large number of individuals in the overall PITCH consortium, because breakthrough infection during this time (pre-Omicron) was still relatively rare. We used the maximum sample size available for this case-control study for each assay type, however, the small number of cases means there was inadequate power to conduct a fully adjusted analysis of the effect of humoral and cellular responses on odds of vaccine breakthrough using logistic regression. This means there may be some confounding in our analysis, for example, due to previous infection. However, non-mechanistic CoP are still useful. There are unfortunately no further stored PBMC samples available for analysis. (ii) Sequence information was not available so we were unable to confirm whether these breakthrough infections were attributable to the Delta variant. However, during the period in which the infections occurred, Delta was the predominant variant in the UK, causing ≥89.9% of genotyped infections throughout this time. This approach using the timing of infection to indicate which variant is likely the cause of infection is in line with approaches used in other studies (Collie et al., 2022). (iii) Our study was not designed to explore specific co-morbidities, particularly immunosuppression, as risk factors for breakthrough infection (Green et al., 2022; Selvavinayagam et al., 2022). This is addressed in the subsequent VIBRANT study (https://vibrant-research-study.org/). (iv) All observational studies carry the risk of bias. Adherence to LFD and PCR testing recommendations was not monitored within PITCH, thus there may be some mis-classification of controls. Although serology from later follow up points was used to confirm as far as possible that controls were not infected during the period of interest, up to 60% of vaccinated individuals may not undergo anti-N seroconversion upon infection (Follmann et al., 2022). It is likely there were differences in exposure to SARS-CoV-2 between individuals dependent on their exact HCW role, for example, seeing more than one COVID-19 patient per day has previously been associated with a higher risk of infection among HCWs (Feng et al., 2021); however, this information was not available for us to take in to account in analysis. (v) We did not investigate mucosal immunity in this study, as mucosal samples were not obtained from PITCH participants at that time. Mucosal immune responses do not necessarily correlate with responses in the peripheral circulation (Lim et al., 2022), and may play an important role in protection against SARS-CoV-2 infection, particularly in individuals with hybrid immunity (Russell and Mestecky, 2022). Ongoing PITCH research now incorporates measurement of mucosal immunity. Nonetheless, this study has several strengths, primarily that the PITCH cohort is very well characterised, and the prospective nature of the study allowed investigation at a pre-specified timepoint reflecting peak vaccine responses, prior to infection.

Questions remain regarding the mechanism of action of these putative T cell CoP. CD8+ T cells may contribute to control of replication after infection, as demonstrated in a rhesus macaque model (Liu et al., 2022). CD4+ T cells are important for providing T cell help to support CD8+ T cells and for generation of Ab responses. There may also be a role for HLA type and which epitopes are immunodominant in different individuals. For example, it has been shown that there is an association between the HLA-DQB1*06 allele and higher anti-RBD IgG titre after vaccination with AZD1222, and that this may play a role in protection from breakthrough infection with ancestral and Alpha strains (Mentzer et al., 2022).

Overall, our study supports a role for T cell responses alongside antibody responses in protection against detectable infection during the period after double vaccination, over which antibody responses have been shown to wane (Levin et al., 2021; Moore et al., 2022; Naaber et al., 2021). Moreover, our results suggest a potential role for variant-specific CD8+ T cell responses. Development of sensitive, quantitative T cell assays that can be used at scale in large cohorts in a range of settings will allow further integrated study of humoral and cellular adaptive immunity and inform vaccine surveillance and effectiveness monitoring.

## METHODS

### Study design and sample collection

The PITCH consortium has been described previously (Angyal et al., 2022; Moore et al., 2022; Payne et al., 2021). Briefly, participants in this prospective, observational, cohort study were recruited from five university hospitals in England (Birmingham, Liverpool, Newcastle, Oxford and Sheffield). Recruitment of consenting individuals was by word of mouth, hospital e-mail communications and from hospital-based staff screening programmes for SARS-CoV-2, including HCWs enrolled in the national SIREN study at three sites (Liverpool, Newcastle and Sheffield). Eligible participants were adults aged 18 or over, and currently working as HCWs, including allied support and laboratory staff, or were volunteers linked to the hospital. Individuals were defined as SARS-CoV-2 naive or previously-infected based on documented PCR and/or serology results from local NHS trusts, or the Meso Scale Discovery (MSD) assay S and N antibody results if these data were not available locally, as described previously (Payne et al., 2021).

PITCH is a sub-study of the SIREN study, which was approved by the Berkshire Research Ethics Committee, Health Research 250 Authority (IRAS ID 284460, REC reference 20/SC/0230), with PITCH recognised as a sub-study on 2 December 2020. SIREN is registered with ISRCTN (Trial ID: 252 ISRCTN11041050). Some participants were recruited under aligned study protocols. In Birmingham participants were recruited under the “Determining the immune response to SARS-CoV-2 infection in convalescent health care workers” (COCO) study (IRAS ID: 282525). In Liverpool some participants were recruited under the “Human immune responses to acute virus infections” Study (16/NW/0170), approved by North West - Liverpool Central Research Ethics Committee on 8 March 2016, and amended on 14th September 2020 and 4th May 2021. In Oxford, participants were recruited under the GI Biobank Study 16/YH/0247, approved by the research ethics committee (REC) at Yorkshire & The Humber - Sheffield Research Ethics Committee on 29 July 2016, which has been amended for this purpose on 8 June 2020. In Sheffield, participants were recruited under the Observational Biobanking study STHObs (18/YH/0441), which was amended for this study on 10 September 2020. The study was conducted in compliance with all relevant ethical regulations for work with human participants, and according to the principles of the Declaration of Helsinki (2008) and the International Conference on Harmonization (ICH) Good Clinical Practice (GCP) guidelines. Written informed consent was obtained for all participants enrolled in the study.

The current study is nested within PITCH. A vaccine breakthrough case was defined as a self-reported positive PCR or LFD test for SARS-CoV-2 >14 days after second vaccine dose and <7 days after the third vaccine dose. The period of interest for cases was when Delta was the predominant circulating SARS-CoV-2 variant in the UK, defined here as 6^th^ June to 27^th^ November 2021, with 27^th^ November taken as the cut-off date as this was the date of the first reported cases of SARS-CoV-2 Omicron variant in the UK (Department of Health and Social Care, 2021).

Throughout this period ≥89.9% of genotyped infections across England were attributable to the Delta variant (UK Health Security Agency, 2021). During this period all UK HCWs were requested to undertake twice-weekly asymptomatic testing for SARS-CoV-2 using self-swab LFDs which were provided free of charge by workplaces, pharmacies and by postal delivery from the government website. Symptomatic PCR testing was readily available at this time free of charge from hospital trusts and UK community testing programmes. Adherence to this testing was not measured within PITCH, however, PITCH participants are asked about SARS-CoV-2 infections at every clinic visit.

The current study has a case-control design (Supplementary Figure S1). Eligibility criteria were enrolment in PITCH by 30^th^ June 2021, and being double vaccinated by 31^st^ July 2021. For correlates analysis, all those meeting the criteria of a vaccine breakthrough case and who had been sampled at the V2+28 timepoint were included. Controls were those who had been sampled at the V2+28 timepoint and did not report a positive PCR or LFD by 27^th^ November 2021 or 7 days after the third vaccine dose (V3+7), whichever was earlier, and who did not show detectable N seroconversion (defined as anti-N IgG titre over the positivity threshold previously defined using pre-pandemic samples (Angyal et al., 2022), and at least double the individual’s baseline value) in available follow-up data during this time (Supplementary Table S1, S2).

A subset of cases was selected on the basis of sample availability for further analysis using ICS and memory B cell FluoroSpot. Controls were matched with these cases for each assay, with matching based on age, sex, previous infection status at enrolment, vaccine manufacturer and dose interval (Supplementary Table S3). Cases in this matched analysis had been enrolled at multiple sites, whereas controls had all been enrolled at the Oxford site. All experiments for this subset analysis were conducted at the Oxford site.

Participants were sampled for the current study between 4^th^ January 2021 and 13^th^ August 2021, approximately 28 days after their second vaccine dose (median 28 days, IQR 26-33 days). PBMC, plasma and serum were separated and cryopreserved. The majority of participants had been investigated for previous reports of the PITCH cohort (Angyal et al., 2022; Moore et al., 2022; Payne et al., 2021; Skelly et al., 2021).

### Demographic analysis

Demographic characteristics of vaccine breakthrough cases compared to the PITCH cohort were initially analysed by univariable logistic regression to estimate the crude odds ratio for vaccine breakthrough for each variable. A likelihood ratio test was conducted with the null hypothesis of no association between vaccine breakthrough and each variable. An adjusted model was derived by backwards stepwise regression, with age and sex included as *a priori* confounders. Variables were removed from the model sequentially, beginning with those with the least effect in univariable regression. A likelihood ratio test was conducted after each removal, and only those variables with evidence of association with vaccine breakthrough (p<0.05) were retained in the model. Analysis was performed using R version 4.1.2 (The R Foundation for Statistical Computing, Vienna, Austria, 2021. URL https://www.R-project.org/). Confidence intervals were derived using the confint package.

### *T cell IFN*γ *ELISpot assay*

IFNγ ELISpot assays were performed using the Human IFNγ ELISpot Basic kit (Mabtech 3420-2A), according to the PITCH ELISpot Standard Operating Procedure as previously published (Angyal et al., 2022). Cryopreserved PBMC were thawed in R10 media (RPMI 1640 with 10% (v/v) Fetal Bovine Serum, 2mM L-Glutamine and 1mM Penicillin/Streptomycin (all Sigma)) and rested for 3-6 hours in a humidified incubator at 37°C, 5% CO_2_. MultiScreen-IP filter plates (Millipore) were coated with 10μg/ml capture antibody (clone 1-D1K, Mabtech) for 3-8 hours and blocked with R10 or Rab10 (RPMI 1640 with 2mM L-Glutamine and 1mM Penicillin/Streptomycin supplemented with 10% (v/v) Human AB Serum (Sigma)) for 1-8 hours at room temperature (RT). PBMC were resuspended in R10 or Rab10, plated in triplicate at 2×10^5^ cells/well and stimulated as follows. PBMC were stimulated with overlapping peptide pools (18-mers with 10 amino acid overlap, Mimotopes) spanning the S or membrane (M) and N SARS-CoV-2 proteins at a final concentration of 2μg/ml for 16-18 hours in a humidified incubator at 37°C, 5% CO_2_. For selected individuals, pools spanning S protein of the Delta variant were included. Pools consisting of CMV, EBV and influenza peptides at a final concentration of 2μg/ml (CEF; Proimmune) and concanavalin A or phytohemagglutinin L (PHA-L, Sigma) were used as positive controls. DMSO was used as the negative control at an equivalent concentration to the peptides. After the incubation period and all subsequent steps plates were washed with PBS supplemented with 0.05% (v/v) Tween20 (Sigma). Detection antibody (clone 7-B6-1, Mabtech) was added and incubated for 2-4 hours at RT. Plates were washed and streptavidin-ALP (Mabtech) added for 1-2 hours at RT, before washing again and addition of 1-step NBT/BCIP substrate solution (Thermo Scientific) for 5-7 minutes at RT. Colour development was stopped by washing with tap water. Air-dried plates were scanned and analysed with the AID Classic ELISpot reader (software version 8.0, Autoimmune Diagnostika GmbH, Germany) or ImmunoSpot® S6 Alfa Analyser (Cellular Technology Limited LLC, Germany). Antigen-specific responses were calculated by subtracting the mean number of spots in the DMSO wells from the test wells and expressed as spot-forming units (SFU)/10^6^ PBMC. Samples with a mean spot value >50 in the DMSO wells were excluded from analysis.

### MSD IgG binding assay

IgG responses to SARS-CoV-2, SARS-CoV-1, MERS-CoV and seasonal coronaviruses were measured using a multiplexed MSD immunoassay (The V-PLEX COVID-19 Coronavirus Panel 3 (IgG) Kit (cat. no. K15399U) from Meso Scale Discovery, Rockville, MD USA), as reported previously (Moore et al., 2022; Payne et al., 2021). A MULTI-SPOT® 96-well, 10 spot plate was coated with three SARS CoV-2 antigens (S, RBD, N), SARS-CoV-1 and MERS-CoV spike trimers, spike proteins from seasonal human coronaviruses, HCoV-OC43, HCoV-HKU1, HCoV-229E and HCoV-NL63, and bovine serum albumin (negative control). Antigens were spotted at 200-400 μg/mL (MSD® Coronavirus Plate 3). Multiplex MSD assays were performed as per the manufacturer’s instructions. To measure IgG antibodies, 96-well plates were blocked with MSD Blocker A for 30 minutes. Following washing with washing buffer, serum or plasma samples diluted 1:1,000-30,000 in diluent buffer, MSD standard and undiluted internal MSD controls, were added to the wells. After 2-hour incubation and washing, detection antibody (MSD SULFO-TAG(tm) anti-human IgG antibody, 1/200) was added. Following washing, MSD GOLD(tm) read buffer B was added and plates were read using a MESO® SECTOR S 600 reader. The standard curve was established by fitting the signals from the reference standard using a 4-parameter logistic model. Concentrations of samples were determined from the electrochemiluminescence signals by back-fitting to the standard curve and multiplying by the dilution factor. Concentrations are expressed in Arbitrary Units/ml (AU/ml). Thresholds were previously determined for each SARS-CoV-2 antigen based on the mean concentrations measured in 103 pre-pandemic sera + 3 Standard Deviations (Angyal et al., 2022). Thresholds were: S, 1160 AU/ml; RBD, 1169 AU/ml; and N, 3874 AU/ml.

### Memory B cell IgG FluoroSpot assay

In a subset of cases and matched controls for which PBMC were available, B cell memory responses were characterised by IgG FluoroSpot assay, according to methods previously reported (Moore et al., 2022). Cryopreserved PBMC were thawed and rested for 4-6 hours in R10 media, then cultured in R10 supplemented with 2mM Sodium Pyruvate, 2mM MEM Non-Essential Amino Acids, 200nM 2-Mercaptoethanol (all Life Technologies), 1μg/ml R848 and 10ng/ml IL-2 (Mabtech Human memory B cell stimpack) for 68-72 hours in a humidified incubator at 37°C, 5% CO_2_, for polyclonal stimulation. Using the Human IgA/IgG FluoroSpotFLEX kit (Mabtech), stimulated PBMC were then added at 2×10^5^ cells/well to FluoroSpot plates coated with 10μg/ml ancestral and Delta SARS-CoV-2 S glycoprotein, PBS (negative control) or capture mAbs (positive control). Plates were incubated for 18 hours in a humidified incubator at 37°C, 5% CO_2_ and developed according to the manufacturer’s instructions (Mabtech). Analysis was carried out with AID ELISpot software 8.0 (Autoimmun Diagnostika). Samples were tested in duplicate, or triplicate where cell counts permitted. Antigen-specific responses were calculated by subtracting the mean number of spots in the PBS wells from the test wells and expressed as antibody-secreting cells (ASC)/10^6^ PBMC.

### Intracellular cytokine staining assay

In a subset of cases and matched controls for which PBMC were available, T cell responses were characterised further using ICS after stimulation with overlapping SARS-CoV-2 peptide pools, according to methods reported previously (Moore et al., 2022). Cryopreserved PBMC were thawed and rested in R10 media for 4-6 hours, then plated at 1×10^6^ cells/well in a 96-well U-bottom plate together with co-stimulatory molecules anti-CD28 and anti-CD49d (both BD) at 1μg/ml final concentration. Peptide pools spanning ancestral and Delta S proteins were added at 2μg/ml final concentration for each peptide pool. DMSO (Sigma) was used as the negative control at the equivalent concentration to the peptides. As a positive control, cells were stimulated with 1x cell activation cocktail containing phorbol-12-myristate 13-acetate (PMA) at 81µM and ionomycin at 1.3µM final concentration (Biolegend). Cells were then incubated in a humidified incubator at 37°C, 5% CO_2_ for 1 hour before incubating for a further 15 hours in the presence of 5µg/ml Brefeldin A (Biolegend). Flow cytometry staining was then performed as follows. Cell staining buffer (Biolegend) was used for staining and washing before fixation, with 1x Perm/Wash buffer (BD) used after fixation. At the end of the culture period, PBMC were washed once and subsequently stained with near-infrared fixable live/dead stain (Invitrogen), fluorochrome-conjugated primary human-specific antibodies against CD4, CD8 and CD14 (all Biolegend) and human Fc blocking reagent (Miltenyi Biotec) for 20 minutes at 4°C in the dark. PBMC were washed and then fixed and permeabilised in Cytofix/Cytoperm buffer (BD) for 20 minutes at 4°C in the dark. PBMC were then washed followed by staining with primary human-specific antibodies against CD3, IFNγ, TNF (all Biolegend), IL-2 (eBioscience) and human Fc blocking reagent for 20 minutes at 4°C in the dark. PBMC were washed, resuspended in cell staining buffer, and stored at 4°C for up to 6 hours until acquisition. Samples were acquired on a MACSQuant analyser 10 and X (Miltenyi Biotec) and analysis was performed using FlowJo software version 10.8.1 (BD Biosciences). Example gating strategy is shown in Supplementary Figure S4. Details of antibodies used are listed in Supplementary Table S4.

### High-throughput live virus microneutralisation assay

In a subset of cases and controls, high-throughput live virus microneutralisation assays were performed as described previously (Faulkner et al., 2021; Wall et al., 2021). Briefly, Vero E6 cells (Institute Pasteur) (Rihn et al., 2021) at 90-100% confluency in 384-well format were first titrated with varying MOIs of each SARS-CoV-2 variant and varying concentrations of a control monoclonal nanobody in order to normalise for possible replicative differences between variants and select conditions equivalent to wild-type virus. Following this calibration, cells were infected in the presence of serial dilutions of patient serum samples. 24 hrs after infection, cells were fixed with 4% final Formaldehyde, permeabilised with 0.2% TritonX-100, 3% BSA in PBS (v/v), and stained for SARS-CoV-2 N protein using Biotin-labelled-CR3009 antibody produced in-house in conjunction with Alexa488-Streptavidin (Invitrogen S32354) and cellular DNA using DAPI (van den Brink et al., 2005). Whole-well imaging at 5x was carried out using an Opera Phenix (Perkin Elmer) and fluorescent areas and intensity calculated using the Phenix-associated software Harmony (Perkin Elmer). Inhibition was estimated from the measured area of infected cells/total area occupied by all cells. The inhibitory profile of each serum sample was estimated by fitting a 4-parameter dose response curve executed in SciPy. Neutralising antibody titres are reported as the fold-dilution of serum required to inhibit 50% of viral replication (IC50), and are further annotated if they lie above the quantitative (complete inhibition) range (titre >2560), below the quantitative range (<40) but still within the qualitative range (i.e. partial inhibition is observed but a dose-response curve cannot be fit because it does not sufficiently span the IC50), or if they show no inhibition at all. For plotting, complete, weak and no inhibition were assigned 5120, 10 and 5 respectively. Details of viruses used in the study are listed in Supplementary Table S5.

### Statistical analysis

Continuous variables are displayed as median and interquartile range (IQR). Unpaired comparisons between two groups were analysed using the Mann-Whitney U test. Paired comparisons between two groups were analysed by the Wilcoxon matched-pairs signed-rank test. Comparison between four groups was carried out using Fisher’s exact test and post-hoc paired Fisher’s exact tests adjusted with the Bonferroni correction. P values are two-tailed. Statistical analyses were performed using R version 4.1.2 (The R Foundation for Statistical Computing, Vienna, Austria, 2021. URL https://www.R-project.org/) <u>and GraphPad Prism 9.4.0.</u>

## Data Availability

All data produced in the present study are available upon reasonable request to the authors

## ACKNOWLEDGEMENTS

We are grateful to all the volunteers who participated in the PITCH study. For the Birmingham participants, the study was carried out at the National Institute for Health Research (NIHR)/Wellcome Trust Birmingham Clinical Research Facility. Laboratory studies were undertaken by the Clinical Immunology Service, University of Birmingham. We thank Prof. Wendy Barclay of Imperial College and the wider Genotype to Phenotype consortium for the Delta strain used for the live virus microneutralisation assay in this study.

## AUTHOR CONTRIBUTIONS

Conceptualization, S.J.D, P.K, T.L, A.R, C.J.A.D, L.T, T.I.d.S, M.C, S.H, V.H; Methodology, S.J.D, P.K, T.L, B.K, S.L, E.J.C, R.B, A.R, C.J.A.D, L.T, T.I.d.S, M.C; Formal analysis, I.N, M.A; Investigation, I.N, M.A, S.L, P.A, M.Y.W, E.J.C, L.S, S.D, M.P, T.A.H.N, R.P.P, E.B, C.J.A.D, A.R, R.B; Resources, A.B, A.D.O; Data Curation, S.C.M, A.S.D; Writing – Original Draft, I.N; Writing – Review & Editing, S.J.D, P.K, T.L, T.I.d.S, L.T., E.J.C, S.L, M.A; Visualization, I.N; Supervision, S.J.D, P.K, T.L, B.K, M.C, T.I.d.S, L.T, A.T; Project Administration, A.S.D; Funding Acquisition, P.K., S.J.D., L.T., T.I.d.S, C.J.A.D., A.R., S.H., V.H.

