## Supplementary information for "CD4+ and CD8+ T cell and antibody correlates of protection against Delta vaccine breakthrough infection: A nested case-control study within the PITCH study"

### Supplementary Tables S1-S5

**Table S1. Demographic characteristics of vaccine breakthrough cases compared to controls included in main correlates analysis.**

| | Cases (%) | Controls (%) |
|---|---|---|
| **Total N** | 32 | 247 |
| **Age (years)** | | |
| Median | 43 | 43 |
| Interquartile range | 31.5-49 | 34-53.5 |
| Range | 22-72 | 22-71 |
| **Sex** | | |
| Female | 26 (81.25%) | 180 (72.9%) |
| Male | 6 (18.75%) | 67 (27.1%) |
| **Ethnicity (self-reported)** | | |
| White | 26 (81.3%) | 175 (70.9%) |
| Asian | 1 (3.1%) | 20 (8.1%) |
| Other | 1 (3.1%) | 11 (4.5%) |
| Unreported | 4 (12.5%) | 41 (16.6%) |
| **BMI (kg/$m^2$)** | | |
| Not obese (<30) | 11 (34.4%) | 98 (39.7%) |
| Obese (≥30) | 2 (6.3%) | 9 (3.6%) |
| Unreported | 19 (59.4%) | 140 (56.7%) |
| **Vaccine regimen** | | |
| AZD1222 | 9 (28.1%) | 32 (13.0%) |
| BNT162b2 (short interval[1]) | 2 (6.3%) | 58 (23.5%) |
| BNT162b2 (long interval[2]) | 21 (65.6%) | 157 (63.6%) |
| **Infection history** | | |
| Naïve | 26 (81.25) | 147 (59.5%) |
| Convalescent | 6 (18.75%) | 100 (40.5%) |

[1]Short interval represents 2-5 weeks between first and second dose.
[2]Long interval represents 6-14 weeks between first and second dose.

**Table S2. Number and key characteristics of participants sampled for each assay.**

| Immune parameter | Assay | Group | Number of individuals | |
|---|---|---|---|---|
| | | | Cases (total N=32) | Controls (total N=247) |
| **IgG binding** | **Meso Scale Discovery assay (MSD)** | Total | **32** | **200** |
| | | AZ/Naive | 7 | 23 |
| | | AZ/Convalescent | 2 | 9 |
| | | Pfizer/Naive | 19 | 100 |
| | | Pfizer/Convalescent | 4 | 68 |
| **IFNγ response** | **T cell ELISpot (WT peptide pools)** | Total | **24** | **191** |
| | | AZ/Naive | 5 | 10 |
| | | AZ/Convalescent | 0 | 1 |
| | | Pfizer/Naive | 17 | 104 |
| | | Pfizer/Convalescent | 2 | 76 |
| | **T cell ELISpot (Delta peptide pools)** | Total | **9** | **25** |
| | | AZ/Naive | 5 | 5 |
| | | AZ/Convalescent | 0 | 0 |
| | | Pfizer/Naive | 3 | 13 |
| | | Pfizer/Convalescent | 1 | 7 |
| **Neutralising antibody** | **Live virus microneutralisation assay** | Total | **21** | **41** |
| | | AZ/Naive | 3 | 7 |
| | | AZ/Convalescent | 1 | 0 |
| | | Pfizer/Naive | 14 | 19 |
| | | Pfizer/Convalescent | 3 | 15 |
| **B cell memory** | **B cell FluoroSpot** | Total | **10** | **11** |
| | | AZ/Naive | 2 | 2 |
| | | AZ/Convalescent | 0 | 0 |
| | | Pfizer/Naive | 8 | 9 |
| | | Pfizer/Convalescent | 0 | 0 |
| **CD4+ and CD8+ T cell response** | **Intracellular cytokine staining (ICS)** | Total | **12** | **12** |
| | | AZ/Naive | 2 | 2 |
| | | AZ/Convalescent | 0 | 0 |
| | | Pfizer/Naive | 10 | 10 |
| | | Pfizer/Convalescent | 0 | 0 |

**Table S3. Demographic characteristics of vaccine breakthrough cases compared to matched controls included in B cell FluoroSpot and intracellular cytokine staining assays.**
All individuals were infection-naïve before vaccination.

| | Cases | Controls |
|---|---|---|
| **B cell FluoroSpot assay** | | |
| **Total N** | 10 | 11 |
| **Age (years)** | | |
| Median | 45 | 42 |
| Interquartile range | 39.8-49 | 40-51.5 |
| Range | 28-63 | 22-66 |
| **Sex** | | |
| Female | 9 | 9 |
| Male | 1 | 2 |
| **Vaccine regimen** | | |
| AZD1222 | 2 | 2 |
| BNT162b2 | 8 | 9 |
| **Intracellular cytokine staining (ICS) assay** | | |
| **Total N** | 12 | 12 |
| **Age (years)** | | |
| Median | 40.5 | 41.5 |
| Interquartile range | 31.8-51 | 30-47.8 |
| Range | 24-72 | 22-66 |
| **Sex** | | |
| Female | 11 | 11 |
| Male | 1 | 1 |
| **Vaccine regimen** | | |
| AZD1222 | 2 | 2 |
| BNT162b2 | 10 | 10 |

**Table S4. Antibodies used for intracellular cytokine staining assay.**

| Marker name | Fluorochrome | Clone | Species reactivity | Host species | Isotype | Manufacturer | Catalogue number | Dilution used |
|---|---|---|---|---|---|---|---|---|
| CD3 | PerCP | UCHT1 | Human | Mouse | IgG1,κ | Biolegend | 300428 | 100 |
| CD4 | APC | RPA-T4 | Human | Mouse | IgG1,κ | Biolegend | 300514 | 200 |
| CD8 | BV510 | RPA-T8 | Human | Mouse | IgG1,κ | Biolegend | 301048 | 600 |
| CD14 | APC-Fire750 | M5E2 | Human | Mouse | IgG2a,κ | Biolegend | 301854 | 200 |
| IFNγ | PE | 4S.B3 | Human | Mouse | IgG1,κ | Biolegend | 502508 | 50 |
| TNF | FITC | MAb11 | Human | Mouse | IgG1,κ | Biolegend | 502906 | 40 |
| IL-2 | PE-Cy7 | MQ1-17H12 | Human | Rat | IgG2a,κ | eBioscience | 25-7029-41 | 100 |

**Table S5. Details of viruses used for live virus microneutralisation assay.**

| Lineage | Mutations present in Spike | Source |
|---|---|---|
| Ancestral [hCoV19/England/02/2020] | | Public Health England, UK |
| Delta<br>[MS066352H] | T19R, K77R, G142D, Δ156- 157/R158G, A222V, L452R, T478K, D614G, P681R, D950N | Prof. Wendy Barclay, Imperial College London, London, UK via the Genotype-to-Phenotype National Virology Consortium (G2P-UK) |

**Supplementary Figures S1-S4**

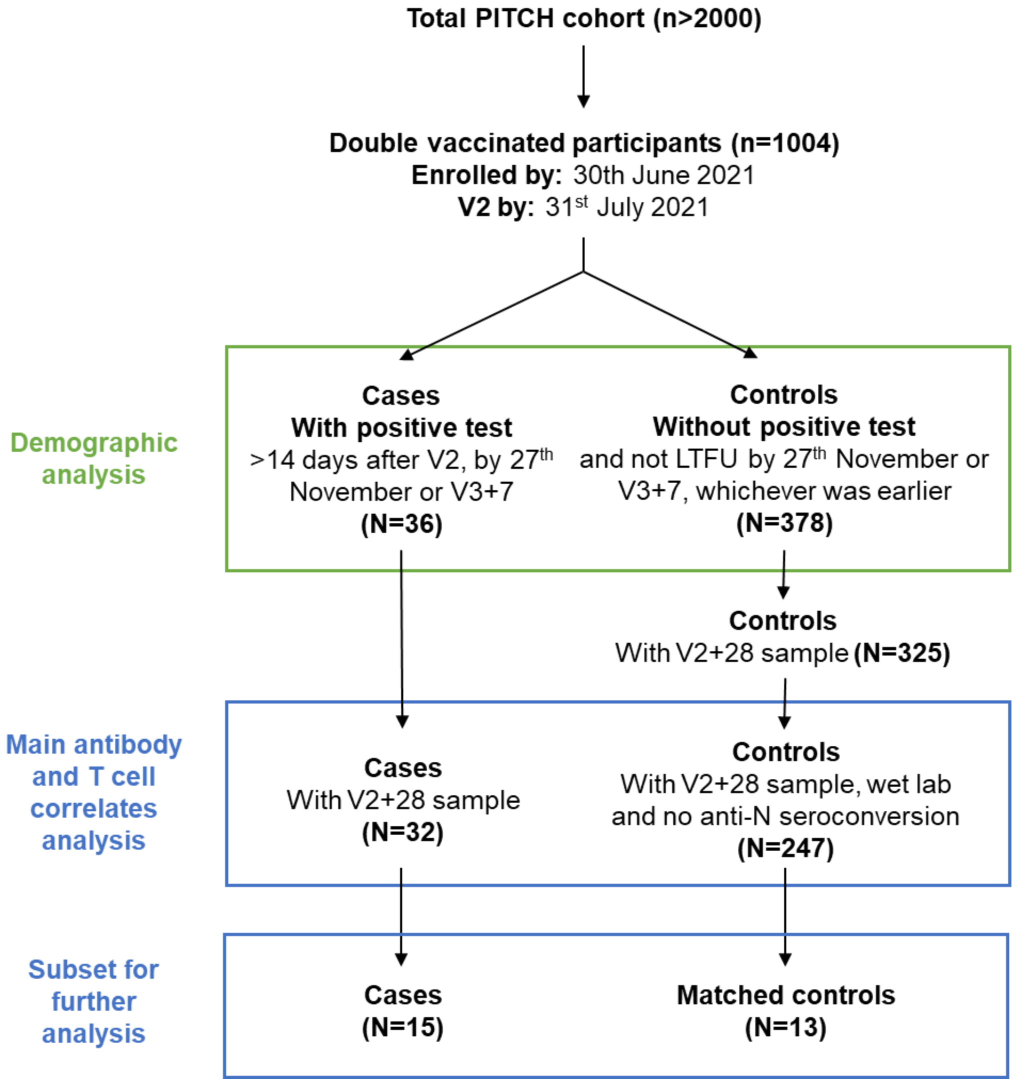


**Figure S1. Flow chart indicating overall design of this study within the PITCH cohort.**
V2 refers to second vaccination. V2+28 refers to 28 days after the second vaccine dose. V3+7 refers to 7 days after the third vaccine dose. LTFU refers to lost to follow-up.

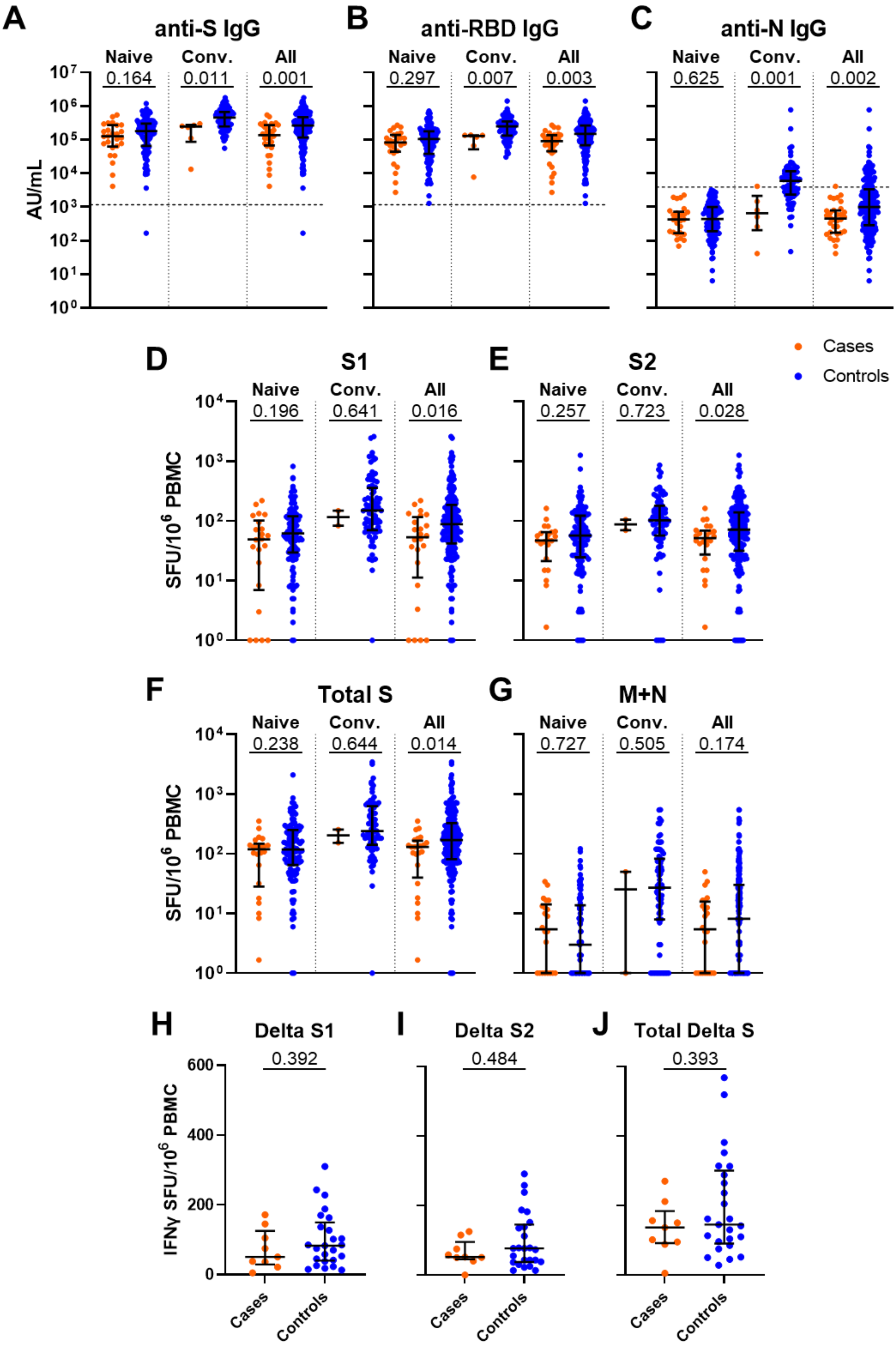


**Figure S2. Comparison of antibody binding and T cell responses between cases and controls according to infection history, and T cell responses to Delta spike peptide pools between cases and controls, at 28 days after second**

**vaccine dose.**
**(A)** Ancestral SARS-CoV-2 spike- (S), **(B)** receptor binding domain- (RBD) and **(C)** nucleoprotein- (N) specific IgG binding titre in cases and controls as measured by MSD, according to infection history. Dashed lines represent threshold for positive response (SARS-CoV-2 S IgG 1160 AU/mL, RDB IgG 1169 AU/mL, N IgG 3874 AU/mL). Naïve represents those with no history of prior infection (cases n=26, controls n=123). Convalescent (Conv) represents those with a history of infection prior to vaccination (cases n=6, controls n=77). All represents summation of Naïve and Conv participants (cases n=32, controls n=200). **(D)** T cell responses to peptide pools representing ancestral SARS-CoV-2 S1, **(E)** S2, **(F)** total S (summation of S1 and S2 responses) and **(G)** membrane (M) and nucleoprotein (N) in cases and controls as measured by IFNγ ELISpot assay, according to infection history. Naïve cases n=22, controls n=114. Conv cases n=2, controls n=77. All cases n=24, controls n=191. Values of 0 are replaced with 1 for representation on the logarithmic scale. **(H)** T cell responses to peptide pools representing Delta (B1.617.2) SARS-CoV-2 S1, **(I)** S2 and **(J)** Total Delta S (summation or Delta S1 and S2 responses) in a subset of cases (n=9) and controls (n=25), as measured by IFNγ ELISpot assay. Orange circles represent cases, blue circles represent controls. Bars represent median of each group. Error bars represent interquartile range. Two-tailed p-values derived from Mann-Whitney U tests shown above linking lines.

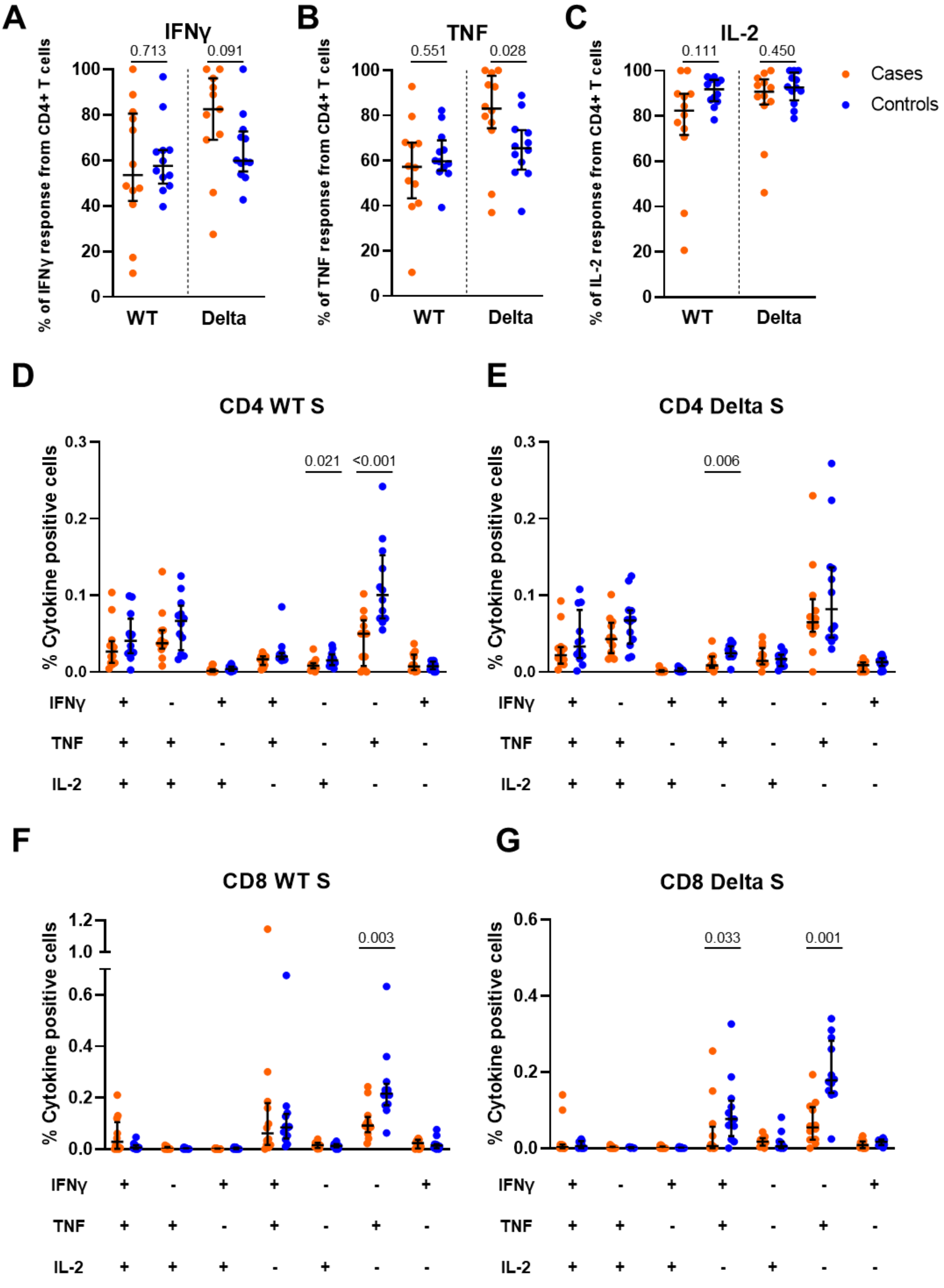


**Figure S3. Comparison of contribution of CD4+ T cell populations to cytokine responses and CD4+ and CD8+ T cell polyfunctionality between cases and matched controls at 28 days after second vaccine dose.**
**(A)** T cell populations responsible for expression of IFNγ, **(B)** TNF and **(C)** IL-2,

calculated by dividing the number of CD4+ cells expressing that cytokine after background subtraction, divided by the total number of CD4+ and CD8+ cells expressing that cytokine after background subtraction. **(D)** T cell polyfunctionality as assessed by combination of expression of IFNγ, IL-2 and TNF cytokines in CD4+ cells in response to ancestral (WT) spike, **(E)** CD4+ cells in response to Delta spike, **(F)** CD8+ cells in response to WT spike and **(G)** CD8+ cells in response to Delta spike peptide pools. Orange circles represent cases, blue circles represent controls. Bars represent median of each group. Error bars represent interquartile range. Two-tailed p-values derived from Mann-Whitney U tests shown above linking lines. For clarity, only p-values <0.05 shown in (D-G).

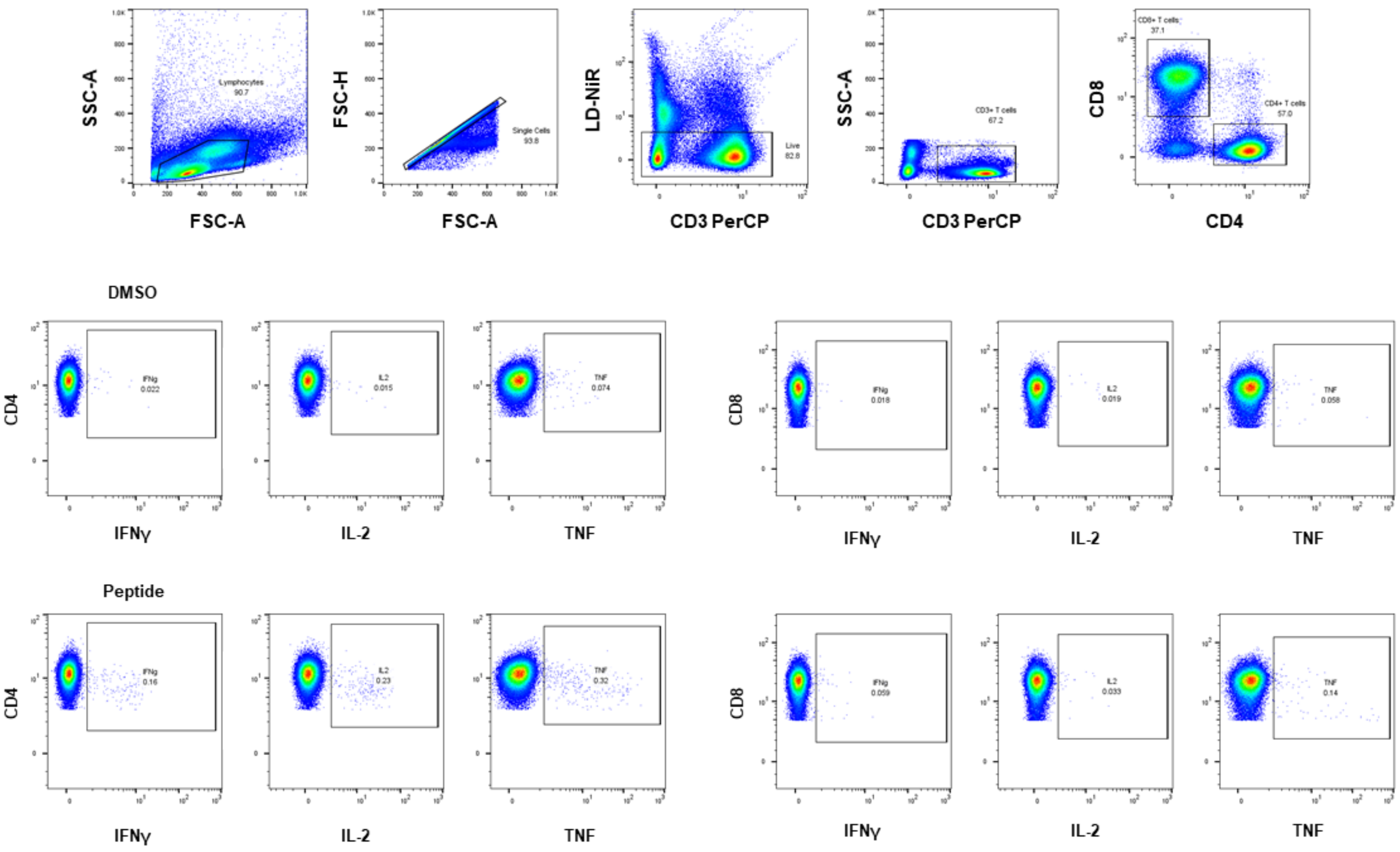


**Figure S4. Gating strategy for intracellular cytokine staining (ICS) assay.** Lymphocytes were gated using forward scatter (FSC)-area (A) and side scatter (SSC)-A. FSC-A and FSC-height (H) were then used to identify single cells. Live CD3+ T cells were gated by first excluding dead cells on the basis of exclusion of Live Dead stain (LD-NiR) and then gating on positivity for CD3 PerCP. CD4+ and CD8+ subsets were identified based on staining for CD4 APC and CD8 BV510. Expression of IFNγ, IL-2 and TNF was identified in the CD4+CD8- and CD4-CD8+ gates. Representative gating shown for one sample for the negative control (DMSO) and ancestral S (peptide) stimulation conditions.

**Secondary Author List**

Crick COVID Immunity Pipeline consortium authors, all affiliated with The Francis Crick Institute, London, UK:

Titilayo Abiola, Janet Abreu, Lorin Adams, Ana Agau-Doce, Karen Ambrose, Neil Bailey, Philip Bawumia, Annabel Borg, Mercedes Cabrera Jarana, Simon Caidan, Marie Caulfield, Bobbi Clayton, Ilenia D'Angelo, Giulia Dowgier, Steve Gamblin, Sonia Gandhi, Mike Gavrielides, Ruth Harvey, Lou Herman, Agnieszka Hobbs, Philip Hobson, Jag Kandasamy, Svend Kjaer, Murad Miah, Mauro Miranda, Sina Namjou, Nicola O'Reilly, Meghan Poulten, Martina Ragno, Mahbubur Rahman, Andrew Riddell, Chloë Roustan, Emma Russell, Samuel Sade, Sandeep Sandhar, Chelsea Sawyer, Vanessa Silva, Callie Smith, Amy Strange, Charles Swanton, Tom Taylor, Rachel Ulferts, Scott Warchal.

PITCH Consortium authors:

| Full Name | Institution |
|---|---|
| Jenna Ablott | Sheffield Teaching Hospitals NHS Foundation Trust |
| Sandra Adele | University of Oxford |
| Zahra Ahmed | University of Birmingham |
| Saly Al-Taei | University of Birmingham |
| Ali Amini | University of Oxford |
| Adrienn Angyal | University of Sheffield |
| M. Azim Ansari | University of Oxford |
| Rachel Anslow | University of Oxford |
| Ana Atti | UK Health Security Agency |
| James Austin | University of Liverpool |
| Angela Bailey | Newcastle upon Tyne Hospitals NHS Foundation Trust |
| Natalie A. Barratt | University of Sheffield |
| Martin Bayley | University of Sheffield |
| Sagida Bibi | University of Oxford |
| Lucy H. Booth | University of Cambridge |
| Alice Bridges-Webb | University of Oxford |
| Rebecca Brown | University of Sheffield |
| Holly Caborn | Sheffield Teaching Hospitals NHS Foundation Trust |
| Jeremy Chalk | University of Oxford |
| Anu Chawla | Liverpool University Hospitals NHS Foundation Trust |
| Elizabeth Clutterbuck | University of Oxford |
| Christopher P. Conlon | University of Oxford |
| Andrew Cross | Liverpool University Hospitals NHS Foundation Trust |
| Debbie Cross | University of Oxford |
| Sophie Davies | University of Oxford |
| Catherine de Lara | University of Oxford |
| Wanwisa Dejnirattisai | University of Oxford |

| | |
|---|---|
| Christina Dold | University of Oxford |
| Thomas M. Drake | University of Edinburgh |
| Elena Efstathiou | University of Birmingham |
| David Eyre | University of Oxford |
| Alex Fairman | University of Sheffield |
| Sian Faustini | University of Birmingham |
| Andrew Filby | Newcastle University |
| Sarah Foulkes | UK Health Security Agency |
| John Frater | University of Oxford |
| Lisa Frending | University of Oxford |
| Oliver Galgut | University of Birmingham |
| Siobhan Gardiner | University of Oxford |
| Philip Goulder | University of Oxford |
| Jessica Gregory | Sheffield Teaching Hospitals NHS Foundation Trust |
| Irina Grouneva | University of Sheffield |
| Lotta Gustafsson | Sheffield Teaching Hospitals NHS Foundation Trust |
| Carl-Philipp Hackstein | University of Oxford |
| Callum Halstead | University of Oxford |
| Sophie Hambleton | Newcastle University |
| Muzlifah Haniffa | Newcastle University |
| Helen Hanson | Newcastle upon Tyne Hospitals NHS Foundation Trust |
| Alexander Hargreaves | University of Oxford |
| Kate Harrington | Sheffield Teaching Hospitals NHS Foundation Trust |
| Jenny Haworth | Newcastle upon Tyne Hospitals NHS Foundation Trust |
| Carole Hays | Newcastle upon Tyne Hospitals NHS Foundation Trust |
| Phoebe Hazenberg | Newcastle upon Tyne Hospitals NHS Foundation Trust |
| Luisa M. Hering | University of Liverpool |
| Emily C. Horner | University of Cambridge |
| Hailey Hornsby | University of Sheffield |
| Fatima Mariam Ilyas | Sheffield Teaching Hospitals NHS Foundation Trust |
| Jasmin Islam | UK Health Security Agency |
| Anni Jämsén | University of Oxford |
| Katie Jeffery | University of Oxford |
| Sile Johnson | University of Oxford |
| Geraldine Jones | Newcastle upon Tyne Hospitals NHS Foundation Trust |
| Mwila Kasanyinga | University of Oxford |
| Sinead Kelly | Newcastle upon Tyne Hospitals NHS Foundation Trust |
| Maqsood Khan | Sheffield Teaching Hospitals NHS Foundation Trust |
| Jon Kilby | University of Sheffield |
| Rosemary Kirk | Sheffield Teaching Hospitals NHS Foundation Trust |
| Allan Lawrie | University of Sheffield |
| Lauren Lett | University of Liverpool |
| Chang Liu | University of Oxford |
| Alison Lye | Sheffield Teaching Hospitals NHS Foundation Trust |
| Tom Malone | University of Oxford |
| Spyridoula Marinou | University of Oxford |

| | |
|---|---|
| Chloe Matthewman | Sheffield Teaching Hospitals NHS Foundation Trust |
| Philippa C. Matthews | Francis Crick Institute |
| David McDonald | Newcastle University |
| Jessica McNeill | Sheffield Teaching Hospitals NHS Foundation Trust |
| Gracie Mead | University of Oxford |
| Naomi Meardon | Sheffield Teaching Hospitals NHS Foundation Trust |
| Alexander J. Mentzer | University of Oxford |
| Shagun Misra | Sheffield Teaching Hospitals NHS Foundation Trust |
| Juthathip Mongkolsapaya | University of Oxford |
| Sam M. Murray | University of Oxford |
| Jeremy M. Nell | Newcastle upon Tyne Hospitals NHS Foundation Trust |
| Alexander R. Nicols | Newcastle University |
| Christopher Norman | Sheffield Teaching Hospitals NHS Foundation Trust |
| Ane Ogbe | University of Oxford |
| Juyeon Park | University of Oxford |
| Brendan A.I. Payne | Newcastle upon Tyne Hospitals NHS Foundation Trust |
| Eloise Phillips | University of Oxford |
| Gareth Platt | University of Liverpool |
| Andrew J. Pollard | University of Oxford |
| Sonia Poolan | Newcastle upon Tyne Hospitals NHS Foundation Trust |
| Nicholas Provine | University of Oxford |
| Chloe Roddis | Sheffield Teaching Hospitals NHS Foundation Trust |
| Stefan Roman | Sheffield Teaching Hospitals NHS Foundation Trust |
| Leigh Romaniuk | Newcastle upon Tyne Hospitals NHS Foundation Trust |
| Patpong Rongkard | University of Oxford |
| Sarah L. Rowland-Jones | University of Sheffield |
| Ayoub Saei | UK Health Security Agency |
| Jose Schutter | University of Sheffield |
| Gavin Screaton | University of Oxford |
| Adrian Shields | University of Birmingham |
| Laura Silva Reyes | University of Oxford |
| Donal Skelly | University of Oxford |
| Nikki Smith | University of Sheffield |
| Jarmila S. Spegarova | Newcastle University |
| Gareth Stephens | Sheffield Teaching Hospitals NHS Foundation Trust |
| Emily Stephenson | Newcastle University |
| Rachel Stimpson | Sheffield Teaching Hospitals NHS Foundation Trust |
| Scarlett Strickland | Sheffield Teaching Hospitals NHS Foundation Trust |
| Krishanthi Subramaniam | University of Liverpool |
| Piyada Supasa | University of Oxford |
| Chloe Tanner | University of Birmingham |
| Lydia J. Taylor | Newcastle University |
| Chitra Tejpal | University of Oxford |
| James E.D. Thaventhiran | University of Cambridge |
| Nicola Tinker | Sheffield Teaching Hospitals NHS Foundation Trust |
| Tom Tipton | University of Oxford |

| | |
|---|---|
| Neal Townsend | University of Birmingham |
| Simon Travis | University of Oxford |
| Nicola Trewick | Newcastle University |
| Stephanie Tucker | Newcastle University |
| Aekkachai Tuekprakhon | University of Oxford |
| Helena Turton | University of Sheffield |
| Jessica K. Tyerman | Newcastle University |
| Zara Valiji | University of Oxford |
| Lisa Watson | Sheffield Teaching Hospitals NHS Foundation Trust |
| Rachel Whitham | Sheffield Teaching Hospitals NHS Foundation Trust |
| Jayne Willson | Sheffield Teaching Hospitals NHS Foundation Trust |
| Barbara Wilson | Newcastle University |
| Joseph D. Wilson | University of Oxford |
| Steven Wood | University of Sheffield |
| Daniel G. Wootton | University of Liverpool |
| Amira A.T. Zawia | Sheffield Teaching Hospitals NHS Foundation Trust |
| Martha Zewdie | University of Oxford |
| Peijun Zhang | University of Sheffield |
